# Deterioration of patients with mental disorders in Denmark coinciding with the invasion of Ukraine

**DOI:** 10.1101/2022.03.26.22272974

**Authors:** Søren Dinesen Østergaard, Christopher Rohde, Oskar Hougaard Jefsen

**Author notes:** <u>**Corresponding author:**</u> Søren Dinesen Østergaard MD, PhD, Department of Affective Disorders, Aarhus University Hospital - Psychiatry, Palle Juul-Jensens Boulevard 175, 8200 Aarhus N Denmark.

## Abstract

The Russian invasion of Ukraine on February 24, 2022 has led to a humanitarian crisis of immense proportions. In turn, this will, inevitably, also lead to a mental health crisis among those affected by the war - either directly or by proxy. The mental health impact of the war in Ukraine is also likely to affect people beyond the borders of the countries directly involved. Here, we investigated whether Danish patients with mental disorders seem to be affected by the war in Ukraine. Specifically, we searched the Electronic Patient Record system of the Psychiatric Services of the Central Denmark Region in the period from January 1, 2022 to March 8, 2022 for clinical notes containing the word *ukrain* (*irrespective of letters and symbols prior to- or following ukrain). We then manually assessed (read) 100 randomly drawn notes containing *ukrain* to determine whether they described worsening of the mental health/symptoms of the patients that was likely attributable to the war in Ukraine. We identified a total of 16,341 adult patients (57% women, mean age 40 years, SD=18) who were registered with at least one clinical note in the Electronic Patient Record system of the Psychiatric Services of the Central Denmark Region in the period from January 1, 2022 to March 8, 2022. During this period, 567,647 clinical notes were entered into the electronic patient record system. Among these, 502 notes from 369 unique patients contained *ukrain* with a sharp rise in incidence following the Russian invasion of Ukraine on February 24, 2022. Among the 100 randomly drawn clinical notes containing *ukrain* (stemming from 94 unique patients), 62 notes from 58 unique patients described worsening of the mental health/symptoms of the patients that seemed attributable to the war in Ukraine. Although causal inference is challenging in this regard, the results of this study suggest that even individuals remotely distanced from the war in Ukraine may be psychologically affected by it - serving as an early warning sign of the potential width of the negative mental health impact from the war, which will, inevitably, hit the Ukrainian population the hardest.

---

The Russian invasion of Ukraine on February 24, 2022, has led to a humanitarian and health crisis of immense proportions.^1^ The loss of lives due to the war is to be counted in thousands and the number of individuals fleeing in millions. In turn, this will, inevitably, also lead to a mental health crisis among those affected by the war – either directly or by proxy.^2^ The mental health impact of the war in Ukraine is likely to affect people beyond the borders of the countries directly involved. Indeed, although causal inference is challenging in this regard, we have previously shown that acts of terrorism and war may affect individuals remotely distanced from these events.^3-5^ Here, we investigated whether the same was the case for the current situation – with emphasis on patients with mental disorders, who are likely to be particularly vulnerable to external stressors such as those imposed by the war in Ukraine.

This study was conducted in accordance with our equivalent assessment of the impact of the COVID-19 pandemic on the mental health of patients with mental disorders.^6^ Specifically, we identified all adult patients (aged ≥18 on January 1, 2022) registered with at least one clinical note in the Electronic Patient Record system of the Psychiatric Services of the Central Denmark Region (CDR)—one of five Danish regions providing inpatient, outpatient and emergency psychiatric care to the approximately 1.3 million inhabitants in the region—in the period from January 1, 2022 to March 8, 2022. Subsequently, we searched for the word “*ukrain*” (*irrespective of letters and symbols prior to- or following “ukrain”) in the clinical notes of the electronic medical records of these patients. We then manually assessed (read) 100 randomly drawn notes containing *ukrain* to determine whether they described worsening of the mental health/symptoms of the patients that was likely attributable to the war in Ukraine (independent assessments performed by OHR and CR with cases of doubt/discrepancies discussed until consensus was reached). Finally, the cases of worsening were labelled according to the dominant type of psychopathology in agreement with Rohde et al.^6^ (see the Supplementary Material for details). The study was approved by the Legal Office of the CDR in agreement with the Danish Health Care Act, §46, Section 2.

A total of 16,341 adult patients (57% women, mean age 40 years, SD=18) were registered with at least one clinical note in the Electronic Patient Record system of the Psychiatric Services of the CDR in the period from January 1, 2022 to March 8, 2022. During this period, 567,647 clinical notes were entered into the electronic patient record system. Among these, 502 notes from 369 unique patients contained *ukrain* with a sharp rise in incidence following the Russian invasion of Ukraine on February 24, 2022 (see Figure 1).

**Figure 1:**
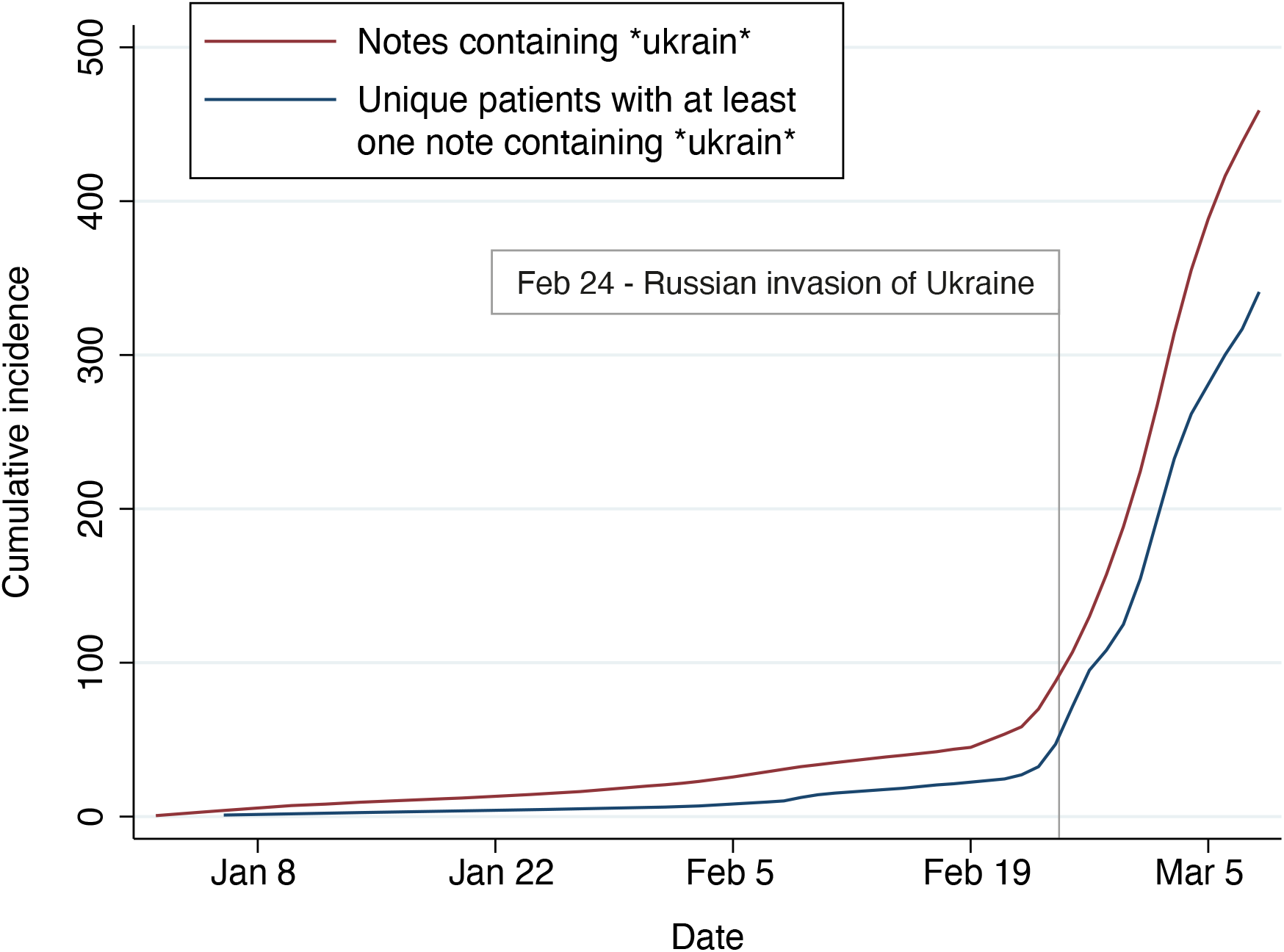
Cumulative incidence of clinical notes containing *ukrain* and patients with at least one clinical note containing *ukrain* over time. Locally weighted smoothing was applied to the lines with a bandwidth of 0.2.

Among the 100 randomly drawn clinical notes containing *ukrain* (stemming from 94 unique patients), 62 notes from 58 unique patients described worsening of the mental health/symptoms of the patients that seemed attributable to the war in Ukraine. The most common type of worsening of psychopathology reported in the notes was anxiety (29%), unspecific stress (21%), reactivation of trauma- or stressor-related symptoms (19%), and delusions/hallucinations (11%), while the most common main diagnoses of the patients were schizophrenia (21%), bipolar disorder (14%) post-traumatic stress disorder (12%) and unipolar depression (16%), respectively. A similar diagnostic pattern was observed among the 369 unique patients with at least one clinical note containing *ukrain* (schizophrenia, 21%; bipolar disorder, 11%; post-traumatic stress disorder, 12%, and unipolar depression, 14%).

The results of this study suggest that the war in Ukraine may have a negative impact on the mental health of patients with mental disorders in Denmark, i.e., making an impact way beyond the borders of Ukraine and its neighboring countries. This is in accordance with our prior studies of the extended effects of terrorism on the mental health of the general Danish population and on refugees in Denmark, respectively,^3-5^ suggesting that the mental health of the Danish population at large is likely to be affected by the current events. Furthermore, given the scale and the media coverage of the war in Ukraine, we find it more likely than unlikely that the observed tendency translates to the populations of other (European) countries as well.

Aside from the obvious fact that establishing causality is inherently challenging in situations where an exposure (in this case a war) cannot be controlled, there are other limitations to this study. First, and relatedly, it cannot be excluded that the patients identified as experiencing worsening of their mental health seemingly related to the war in Ukraine would have felt the same level of distress had Russia not initiated the war. The impression from clinical practice is, however, that we have identified a true signal and not merely a coincidental correlation between the war and patient deterioration. Second, the data stem from everyday clinical practice where the patients with mental disorders were—by no means—systematically assessed for worsening in relation to the war in Ukraine. Third, we employed a quite narrow search focusing exclusively on *ukrain* to obtain high specificity, at the cost of sensitivity. It follows from the two latter limitations that our results should not be interpreted from an absolute perspective (i.e., the number of cases of worsening of psychopathology seemingly related to the war), but rather from a relative and temporal perspective (i.e., the marked increase in the number of cases coinciding with the eruption of war).

In conclusion, the results from this study suggest that even individuals remotely distanced from the war in Ukraine may be psychologically affected by it - serving as an early warning sign of the potential width of the negative mental health impact from the war, which will, inevitably, hit the Ukrainian population the hardest. The mental health- and psychiatric services of the European countries must prepare to meet the increased demand for care as a consequence of the war in Ukraine.

## Data Availability

The data cannot be shared due to restrictions enforced by Danish law for protecting patient privacy. The data is only available for research projects conducted by employees in the Central Denmark Region following approval from the Legal Office under the Central Denmark Region (in accordance with the Danish Health Care Act paragraph 46, Section 2).

## Acknowledgements

The authors are grateful to Bettina Nørremark from Aarhus University Hospital – Psychiatry, for her assistance with data extraction.

## Data availability statement

The data cannot be shared due to restrictions enforced by Danish law for protecting patient privacy. The data is only available for research projects conducted by employees in the Central Denmark Region following approval from the Legal Office under the Central Denmark Region (in accordance with the Danish Health Care Act §46, Section 2).

