## Supplementary Material for "Deterioration of patients with mental disorders in Denmark coinciding with the invasion of Ukraine"

**Labelling of pandemic-related psychopathology**

In agreement with Rohde et al.,^1^ the 100 clinical notes describing psychopathology related to the war in Ukraine were labeled based on the following code:

**1. Substance abuse**

**2. Delusions**

**3. Hallucinations**

**4. Negative symptoms**

**5. Mania/hypomania/mixed state (**”irritable”, ”restless”, overconfident or risky behavior etc.)

**6. Depression** (”sad”, ”unhappy”, ”lonely”, ”down”, ”hopeless”, ”guilty” etc.)

**7. Suicidality/self-harm**

**8. Anxiety (**”worried”, ”anxious”, ”tense”, ”frightened”, ”panicky”, + autonomic symptoms)

**9. Trauma- or stressor-related symptoms** (”alert”, ”vigilant”, “flashbacks”, ”nightmares”, etc.)

**10. Obsessions or compulsions**

**11. Feeding or eating disorders** (anorexia, bulimia, binge eating)

**12. Autism spectrum symptoms** (exacerbated internalizing or externalizing behaviors)

**13. ADHD-related symptoms** (impulsitivy, hyperactivity, attention difficulties)

**14. Disruptive behaviour in children** (antisocial, defiant, oppositional, violent)

**15. Tics (motor/verbal)**

**16. Aggression** (“anger”, “frustration”, “externalizing behaviors”)

**17. Unspecific stress** (”stressed”, ”under pressure”, ”dissatisfaction”/”decreased well-being**”**) not better accounted for by the above symptom domains.

**18. Other symptoms / miscellaneous**

Only one code (the dominant psychopathology) was given per clinical note. Two examples of potentially ambiguous cases are discussed and resolved below:

- Patients with psychotic depression or psychotic mania experiencing delusions or hallucinations likely to be caused by the war in Ukraine were classified as ”2” (delusions) or ”3” (hallucinations), respectively.*

- Patients with any diagnosis (e.g. depression) among whom the war in Ukraine appeared to have been related to self-harm, suicidal thoughts/behavior were classified as ”7” (suicidality/ self-harm).

* This rule was considered to be most informative because the affective aspect would be covered by the patients’ diagnosis, which was also reported in the manuscript.

^1^ Rohde C, Jefsen OH, Nørremark B, et al. Psychiatric symptoms related to the COVID-19 pandemic. Acta Neuropsychiatr. 2020;32:274–276.
